# Altered excitation-inhibition balance in the somatomotor and default mode network in multiple sclerosis

**DOI:** 10.64898/2026.01.14.26344097

**Authors:** Gaia Zin, Guy Nagels, Jeroen Van Schependom, Thanos Manos

## Abstract

**Introduction:** The balance between excitatory and inhibitory (E/I) neural processes is a fundamental principle of brain function, and its disruption has been implicated in the pathophysiology of multiple sclerosis (MS). In vivo assessment of E/I balance has traditionally relied on electrophysiological measures, and despite the abundance of fMRI data on MS, no fMRI-based technique has so far been presented to measure E/I balance in MS.

**Methods:** Recently, a novel MRI-based method has been introduced to estimate E/I balance by integrating functional MRI and diffusion weighted imaging data. We use this approach to study E/I balance in MS at a global (over the whole head) and at the local level of specific resting state networks affected by MS: the somatomotor and Default Mode network (DMN). Furthermore, we perform the analysis using three different atlases: the Schaefer atlas, which is functionally defined, and the Automatic Anatomical Labeling (AAL) and Desikan Killany (DK) atlas, which are defined based on structural features.

**Results:** Our findings reveal a significant alteration in E/I balance within the somatomotor and default mode networks when using the functionally defined Schaefer atlas, suggesting a network-specific dysfunction in MS. We also find that the E/I balance inferred within the somatomotor network correlates with motor fatigue.

**Conclusions:** This study demonstrates a promising framework for investigating E/I balance alterations in neurological disorders and paves the way for validation in larger cohorts.

## 1 INTRODUCTION

Multiple sclerosis (MS) is a chronic neuroinflammatory disease that primarily affects the central nervous system and is the leading cause of non-traumatic neurological disability in young adults [1] and, in particular, women (see e.g., [2]). The pathophysiology of MS is driven by immune-mediated processes that lead to inflammation, demyelination, and neuronal loss (neurodegeneration) [3, 4]. These processes result in a wide range of symptoms, including fatigue and cognitive impairment, see e.g., [5].

A fine balance between excitation and inhibition in the brain, also referred to as Excitation/Inhibition (E/I) balance, is crucial for proper information processing and healthy neuronal function, see e.g., [6, 7]. Disruptions in E/I balance have been observed in various neurological disorders, including schizophrenia [8, 9], Alzheimer’s disease [10, 11], and autism spectrum disorder [12, 13].

In MS, neuronal and synaptic loss could result in a shift in E/I balance [14, 15]. Post-mortem analyses of motor cortex tissue suggest a preferential loss of inhibitory interneurons in the grey matter [16]. Consistent with this, experimental autoimmune encephalomyelitis studies demonstrate a pro-excitatory shift in the motor cortex [17]. Furthermore, Huiskamp et al. [14] observed a loss of both excitatory and inhibitory synapses in post-mortem tissue. These synaptic alterations were subsequently incorporated into a computational model to study large-scale network dynamics in people with multiple sclerosis (pwMS). The authors found that the inhibitory synaptic loss alone impacts the empirical neuronal activity and functional connectivity more profoundly. We also note that in-vivo studies of E/I alterations in MS predominantly rely on electrophysiological modalities like electroencephalography (EEG) and magnetoencephalography (MEG) (see e.g., [18]). However, despite the abundance of fMRI data of pwMS, no fMRI-based study has been used to infer E/I changes in MS.

Recently, there has been growing interest in the interplay between structural and functional connectivity (see e.g., [19]). Structural connectivity can constrain functional dynamics, while functional mechanisms can reshape anatomical pathways through plasticity and neuromodulation [20, 21]. To capture these interactions, Ajilore et al. [22] introduced the concept of a hybrid resting-state structural connectivity (rsSC) as a metric encapsulating the interplay between structural and functional connectivity. The rsSC integrates information from diffusion-weighted imaging (DWI) and resting-state fMRI data. Fortel et al. [23] proposed a similar multimodal rsSC based on a model from statistical mechanics. Through this modeling framework, the entries of the new rsSC are linked to macroscopic E/I dynamics. The authors have used it to study E/I alterations in Alzheimer’s disorder. In particular, female carriers of the apolipoprotein E epsilon 4 exhibited more susceptibility to network disruption, given an increase in excitations, compared to male carriers [24]. Later studies incorporate E/I estimates at both region-of-interest (ROI) and whole-atlas levels into mixed-effects models accounting for sex [25] and disease progression over time [11]. These findings suggest that the rsSC metric reliably captures biologically meaningful differences in E/I balance. Recently, in Manos et al. [26], the authors used such hybrid (structural and functional) connectivity to study whole-brain dynamics and simulated efficiently BOLD signals and functional connectivity with a Kuramoto model. They found that simulations of dynamical models using rsSC to define brain node interactions, instead of classical SC matrices, showed significantly higher correspondence with empirical fMRI data.

This multimodal approach is particularly interesting for MS, where functional brain reorganization likely follows structural damage in the disorder [27]. In particular, we are interested in measuring E/I imbalance both at a global level (whole brain) and at a local level of specific Resting State Networks (RSNs) affected by MS. We focus on two functional networks that play a role in MS: the somatomotor network and the DMN.

The DMN includes areas in the parietal, temporal, and frontal lobes [28]. Bonavita et al. [29] compared functional connectivity in DMN regions in three groups: healthy controls (HC), MS patients with no established cognitive impairment, and cognitively impaired MS patients. They found significantly weaker functional connectivity strength in people with MS (pwMS) compared to HCs, especially in the cingulate cortex. Other studies found DMN changes in functional connectivity strength when comparing different phenotypes of MS [30], and different levels of cognitive function and disease duration [31].

The somatomotor network also plays a key role in MS. In particular, it is associated with fatigue in MS [32], which is one of the most debilitating symptoms reported by patients. Furthermore, maladaptive reorganization of the somatomotor network associates with pain [33, 34]. Radetz et al. [35] observed a lower neurite density in the motor cortex of pwMS, which was correlated with higher excitability thresholds. Furthermore, in an experimental model of autoimmune encephalomyelitis, Potter et al. [17] found increased excitability in specific nodes of the somatomotor network, supporting a pro-excitatory shift in E/I balance within the somatosensory cortex.

In this study, we assessed whether the E/I balance - as captured through multimodal modelling of fMRI and DWI data - can capture differences between people with MS and healthy controls. In addition, we assessed the extent to which this variation depends on the parcellation atlas. Specifically, we employed a functional brain atlas—the Schaefer atlas—which parcellates the brain into regions based on patterns of functional activity, and hence allows for fast identification of well-defined functional networks. To further test our findings and initial intuition regarding the atlas choice, we also included two structural atlases: the AAL and DK atlas, both of which define regions according to anatomical features. By incorporating both functional and structural atlases, we can get insights into how atlas selection influences excitation/inhibition (E/I) estimates and group comparisons.

## 2 MATERIALS AND METHODS

### 2.1 Participants

This study includes data from 12 Healthy Controls (HC) and 24 people with MS (pwMS). All the subjects are female, and the data collection resided within the UZ Brussel. The groups are matched in sex, age, and years of education **Table** 1. The MS group includes only individuals with relapsing-remitting MS, whose diagnosis matches the revised McDonald criteria [36] and have an Expanded Disability Status Score (EDSS) less than 6. Each subject was evaluated with the Fatigue Scale for Motor and Cognitive Functions (FSMC), the Symbol Digit Modalities Test (SDMT), the Controlled Oral Word Association Test (COWAT), and the Brief Visuospatial Memory Test–Revised scores (BVMT-R).

**Table 1.** Description of the clinical parameters of the HC and MS group. We show the mean and standard deviation of clinical parameters for both the HC and MS groups. For EDSS, we present the median and interquartile range (IQR). We applied the Shapiro-Wilk test to assess the normality of age, education, disease duration, EDSS, and cognitive scores in both groups. Most variables did not follow a normal distribution. Therefore, we used the non-parametric Mann-Whitney U test to compare the groups.

|  | HC | pwMS | Group differences<br>(*p*-values) |
| --- | --- | --- | --- |
| N | 12 | 24 | - |
| Sex | F | F | - |
| Age (yrs) | 41±13 | 48±11 | 0.15 |
| Education (yrs) | 14±4 | 13±3 | 0.54 |
| Disease duration (yrs) | - | 15±8 | - |
| EDSS median [IQR] | - | 2.5 [2-3.5] | - |
| **Disability scores** |  |  |  |
| FSMC-motor | 20±7 | 32±9 | 0.0008** |
| FSMC-cognitive | 20±5 | 32±10 | 0.003** |
| SDMT | 52±12 | 50±10 | 1.00 |
| COWAT | 80±20 | 81±26 | 0.75 |
| BVMT-R | 29±4 | 26±6 | 0.16 |

### 2.2 Data acquisition

MRI data were acquired on a 3T Philips Achieva scanner at UZ Brussel. Resting-state fMRI used a 230 × 230 mm field of view, 48 slices, 1.8 × 1.8 × 2.7 mm voxels, flip angle 90°, TR = 3 s, and TE = 5 s. DWI was acquired on the same scanner with 32 non-collinear directions (b = 800 s/mm^2^) and one b0 volume, using a 250 × 250 mm field of view, 70 slices, 0.975 × 0.975 mm in-plane resolution, 2-mm slice thickness, TR = 5133 ms, and TE = 95 ms. Additional T1-weighted images were collected with a 240 × 240 mm field of view, 310 slices, 0.5-mm isotropic voxels, flip angle 8°, TR = 5.19 ms, and TE = 2.30 ms.

### 2.3 Ethics

Patient recruitment took place between 2015 and 2018. All participants provided written informed consent before inclusion. The ethical committees of the National MS Center Melsbroek and UZ Brussel approved the study (Commissie Medische Ethiek UZ Brussel, B.U.N. 143201423263, 2015/11). The study complied with all applicable ethical guidelines for research involving human participants.

### 2.4 DWI processing

We processed the Diffusion Weighted Images (DWI) using MRtrix3 and FSL [37, 38]. The preprocessing includes denoising, corrections for Gibbs ringing artifacts, eddy currents, and inhomogeneity distortions. The non-linear registration with the standard Montreal Neurological Institute (MNI) reference space was performed using ANTs [39]. The tracks are generated with a probabilistic diffusion tensor imaging algorithm, and seeded at the gray matter–white matter interface. After generating the tracks, the structural connectivity is calculated with the ‘tck2connectome’ function of MRtrix3, for each one of the investigated atlases.

### 2.5 fMRI processing

We perform the functional analysis using MATLAB 2024a and SPM12 [40]. Specifically, we apply the lesion segmentation tool from SPM12 to perform lesion growth and filling. We then use the resulting T1 lesion-filled image as a reference for projecting onto the MNI reference space and aligning it with the chosen brain atlas. For the fMRI data, we discard the first ten volumes to minimize startup effects. We apply motion and slice timing corrections to the remaining data. We discard any images that require more than 1 mm of translational or 0.0125 radians of rotational correction to maintain quasi-stationarity [41, 42]. We then linearly register the corrected fMRI images with the lesion-filled T1 image and compute the average signal for each region of interest (ROI). Next, we regress out the confounding effects of motion artifact, white matter and cerebrospinal fluid. After that, we apply bandpass filtering to the fMRI data within the frequency range of 0.009 Hz to 0.08 Hz. We calculate the Pearson correlation coefficient between ROI pairs to assess functional connectivity. Finally, we discard any nonsignificant edges (threshold at *p* = 0.05).

### 2.6 Brain Atlases

In this section, we present the three brain atlases used in our study:

- The Schaefer atlas [43] defines regions of interest (ROIs) from a data-driven approach based on functional MRI recordings. It provides parcellations at multiple resolutions; we used the 100-ROI version. The ROIs follow the Yeo-7 network division [44] and thus map onto seven large-scale resting-state networks (RSNs): visual, somatomotor, dorsal attention, ventral attention (salience), limbic, frontoparietal, and default-mode.
- The Automatic Anatomical Labeling (AAL) atlas [45] comprises 90 cortical and 26 cerebellar ROIs. These ROIs are manually delineated based on sulcal anatomy and then labelled using automated procedures.
- The Desikan–Killiany atlas [46] contains 68 cortical ROIs manually defined using gyral anatomy. It was built from 40 MRI scans originally collected for Alzheimer’s disease research, covering a wide age range (19–86 years) and capturing inter-individual variability relevant to neurodegeneration.

All atlases were obtained from Neuroparc [47], a standardized repository of human brain atlases. Neuroparc supplies atlases aligned to MNI space and organized at three spatial resolutions. While the ROIs of the (functional) Schaefer atlas are divided into RSNs by construction, this is not the case for the other two considered atlases. To align DK ROIs with the somatomotor network and DMN, we follow the partition into RSNs employed by Lee and Frangou [48]. For the AAL atlas, we overlap it with the Yeo-7 parcellation and compute, for each AAL ROI, the proportion of voxels that fall within each Yeo-defined network. We then select, for each RSN, the minimal voxel-percentage threshold that reliably represents that network; the optimal threshold varies across RSNs. We provide a detailed description of this workflow in Section 3 of the Supplementary Material.

### 2.7 Calculation of hybrid rsSC and E/I balance

The rsSC, as proposed by Fortel et al. [23, 49], is a hybrid connectivity combining structural and functional information. It aims to capture excitation-inhibition dynamics at a macroscopic level, especially in cases where regions show strong functional connectivity but lack direct structural links. This approach draws inspiration from the Ising model from statistical mechanics [50]. The Ising model was originally developed to model the (functional) interactions of magnetic dipoles arranged in a lattice. Precisely, it begins with a defined structural layout and infers binary functional states (spins) by minimizing the system’s energy. Researchers have since applied the Ising model to the study of neural dynamics [51, 52]. The rsSC framework follows the inverse direction. It tries to infer the most suitable structural connectivity pattern that explains the observed functional states, a process known as structure-by-function embedding. The full algorithm details are presented in the Supplementary Material, section 1.**Figure** 1 illustrates the schematic of the pipeline, which is run for each of the studied parcelation atlases.

**Figure 1.**
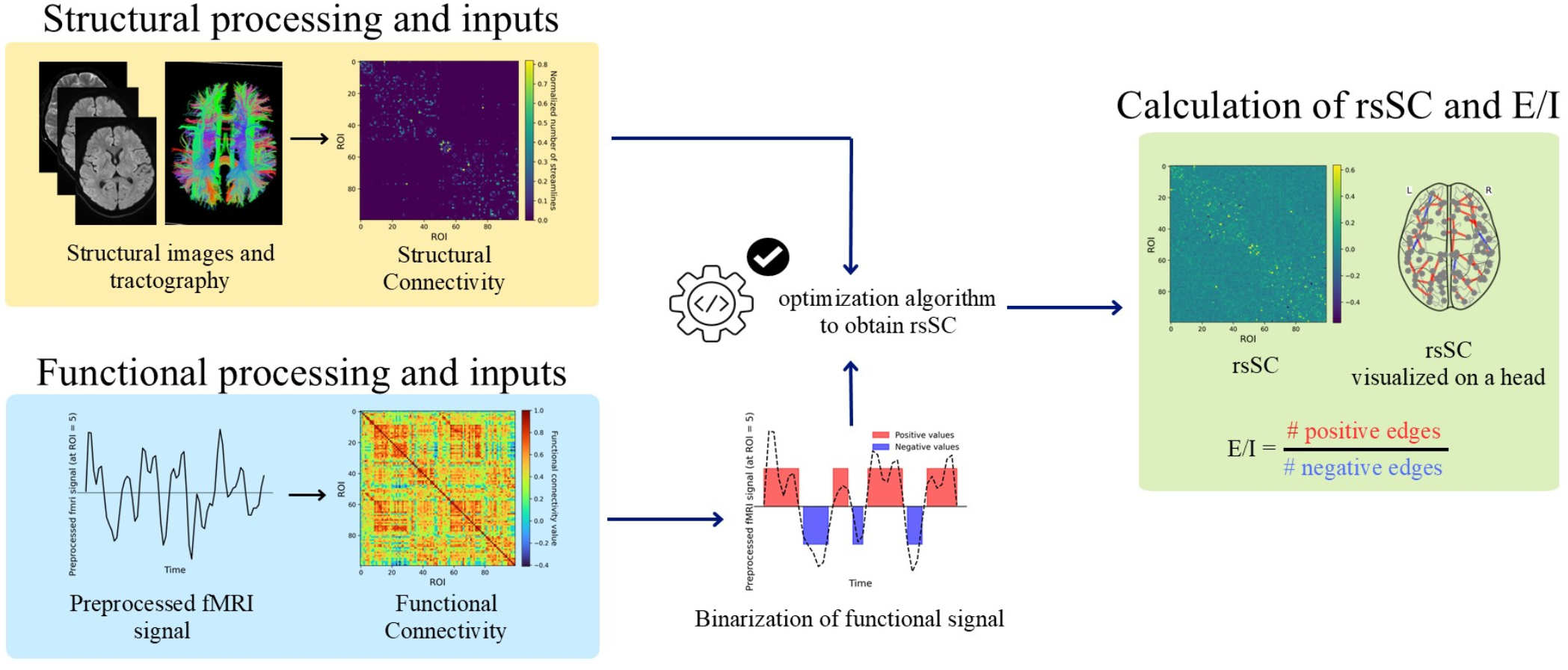
Pipeline to obtain the rsSC and the E/I metric. We depict the pipeline for the calculation of the rsSC and the E/I metric. It takes as input the structural connectivity (in yellow block) and the binarized and preprocessed fMRI signals. The inputs are fed to a structure-by-function embedding, acting as the inverse process to the Ising model, and produce the rsSC. This multimodal matrix contains both positive and negative values. The E/I is calculated as the ratio of the number of positive and the number of negative values in the rsSC.

The resulting connectivity includes both positive and negative values representing macroscopically excitatory and inhibitory dynamics, respectively. Consequently, we calculate the E/I balance metric as the ratio between the number of positive and negative values in the rsSC. The computation is depicted in Eq. (1). Here, *n*_positive_ and *n*_negative_ are the number of positive and negative elements within the rsSC matrix, respectively.

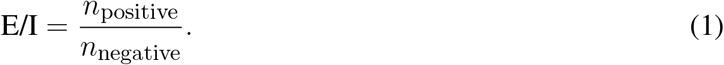

The calculated metric serves as a bridge between microscale neural processes and large-scale network organization, helping to interpret how local activity influences the whole-brain connectivities. We investigate the excitation–inhibition (E/I) balance at global and at local network levels. After computing the E/I balance across all regions of interest (ROIs), we examine it within distinct resting-state networks (RSNs)—specifically, the default-mode network (DMN), the somatomotor network. For each RSN we calculate the E/I balance (EIB) under three conditions:

- **Intra-network:** connections among ROIs that belong to the same network. We obtain this metric from the upper triangle of the sub-matrix comprising only the ROIs of the network.
- **Inter-network:** connections between ROIs inside the network and ROIs outside it. This measure derives from the rectangular sub-matrix whose rows correspond to the network’s ROIs and whose columns correspond to external ROIs.
- **Combined intra- and inter-network:** the union of the two previous sets of connections.

### 2.8 Statistical Analysis

To assess group differences in E/I balance between HC and pwMS, we used the Mann–Whitney U test, a non-parametric method appropriate for small sample sizes. Effect sizes were quantified with Cliff’s δ, a rank-based measure estimating the probability that a value from one group exceeds one from the other. This metric is well-suited for small or unequal groups. To control for multiple comparisons, we applied FDR correction. Observations from different atlases were treated as independent; thus, for each atlas, we corrected across the two RSNs and the three conditions (intra-, inter-, and combined intra–inter-network). After correction, *p <* 0.05 was considered significant.

## 3 RESULTS

### 3.1 Global E/I balance across the whole brain

After computing the rsSC matrices for each atlas (see Supplementary Material, section 2 for visual examples of structural, functional, and hybrid connectivity matrices), we calculated the global excitation/inhibition (E/I) balance (as in Eq. (1)) for both HCs and pwMS using all regions of interest (ROIs) in the hybrid rsSC matrix. **Figure** 2 shows the distribution of global E/I balance, calculated for all three brain parcellations (AAL, DK, and Schaefer). We found no statistically significant differences in E/I balance between groups for any of the atlases. Furthermore, the effect sizes were either negligible or small (see **Table** 2).

**Table 2.** Statistics for the differences in E/I values between HC and pwMS, calculated for ROIs over the whole head. We present the *p*-values from the Mann–Whitney U test and the corresponding Cliff’s delta effect sizes for EIB values in the HC and pwMS groups, computed using the AAL, DK, and Schaefer atlases. We did not find any significant differences in EIB values between healthy controls and individuals with MS across any of the atlases. The associated effect sizes are either small or negligible.

|  | AAL | DK | Schaefer |
| --- | --- | --- | --- |
| $p$-value | 0.46 | 0.63 | 0.89 |
| Cliff’s Delta effect size | 0.15 | 0.099 | -0.032 |

**Figure 2.**
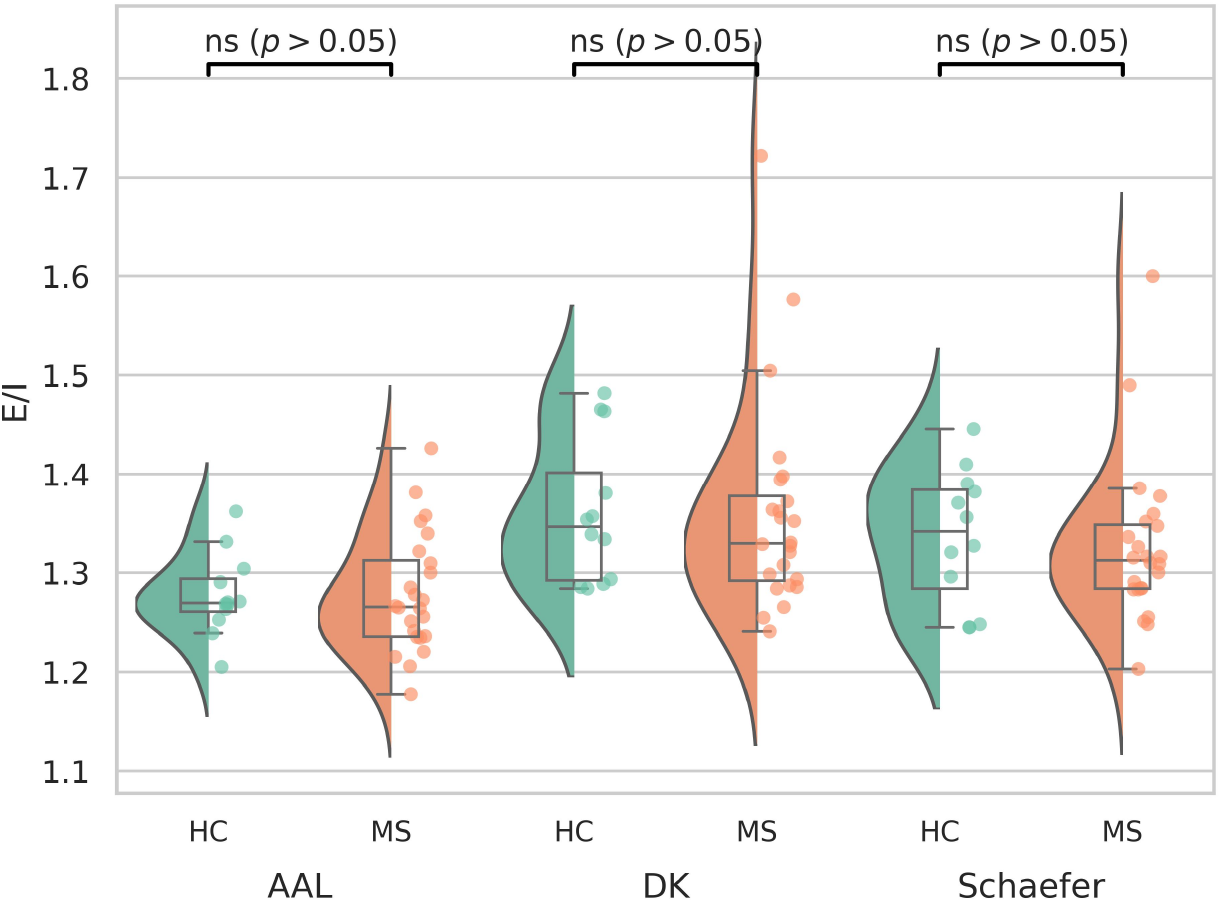
Violin plot of the E/I balance values computed across the whole brain, for HCs and pwMS. We show the violin plots of the E/I distributions computed across all ROIs for the three atlases (AAL, DK, and Scheafer) between the HC (in green) and pwMS (in orange). We do not observe significant differences between E/I balance values of HC and pwMS.

### 3.2 Local E/I balance calculated in the DMN and somatomotor network

Next, we analyzed the E/I balance within the somatomotor network and DMN. For each network, we first looked at intra-network E/I balance (see **Figures** 3(A) and 4(A) respectively). We found that the MS group had significantly lower E/I balance than the HC group in both networks after FDR correction, but only when using the Schaefer atlas (see **Figures** 3(B) and 4(B) respectively). The *p* −value and effect size for these analyses are reported in **Table** 3.

**Table 3.**
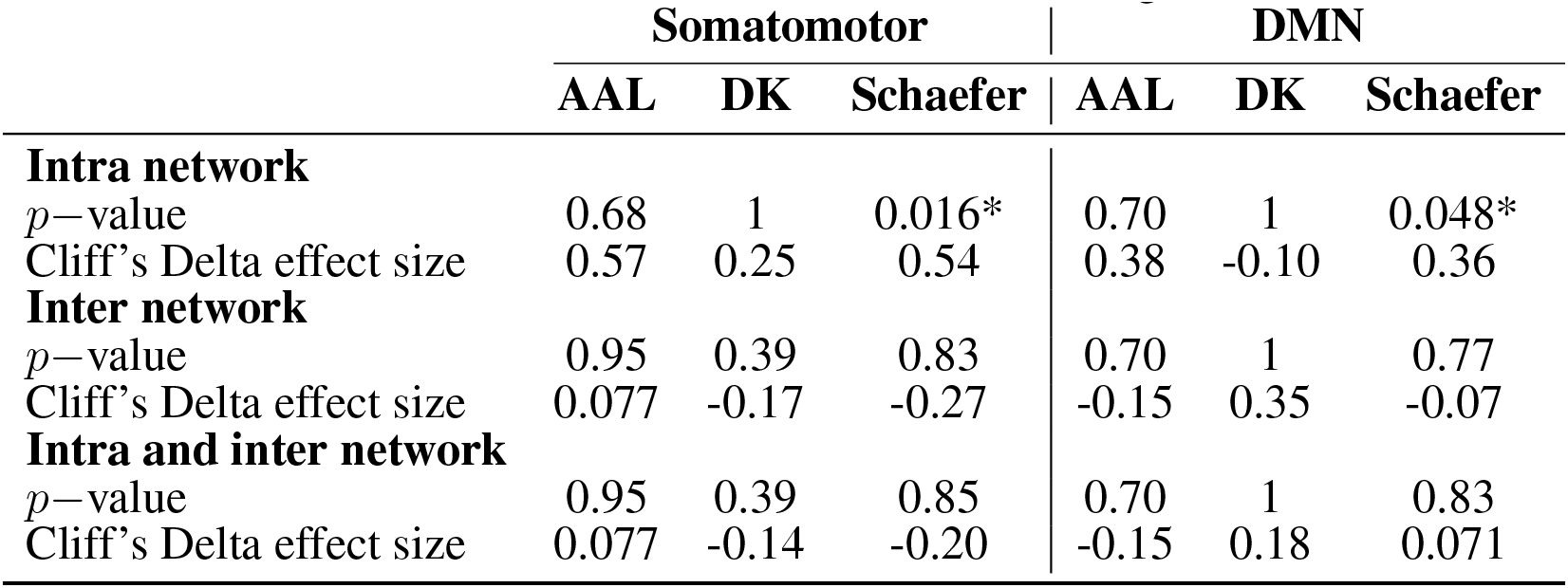
Statistical analysis of EIB values in the somatomotor network and DMN across different brain atlases, comparing HCs and pwMS. EIB values were computed for the somatomotor and DMN network as defined by the AAL, Desikan–Killiany (DK), and Schaefer atlases. For each atlas, intra-network connections, inter-network connections, and combined intra- and inter-network connections were analyzed. Significant group differences were observed in intra-network EIB values for the Schaefer atlas for both the somatomotor and DMN. The associated effect sizes are large and medium.

**Figure 3.**
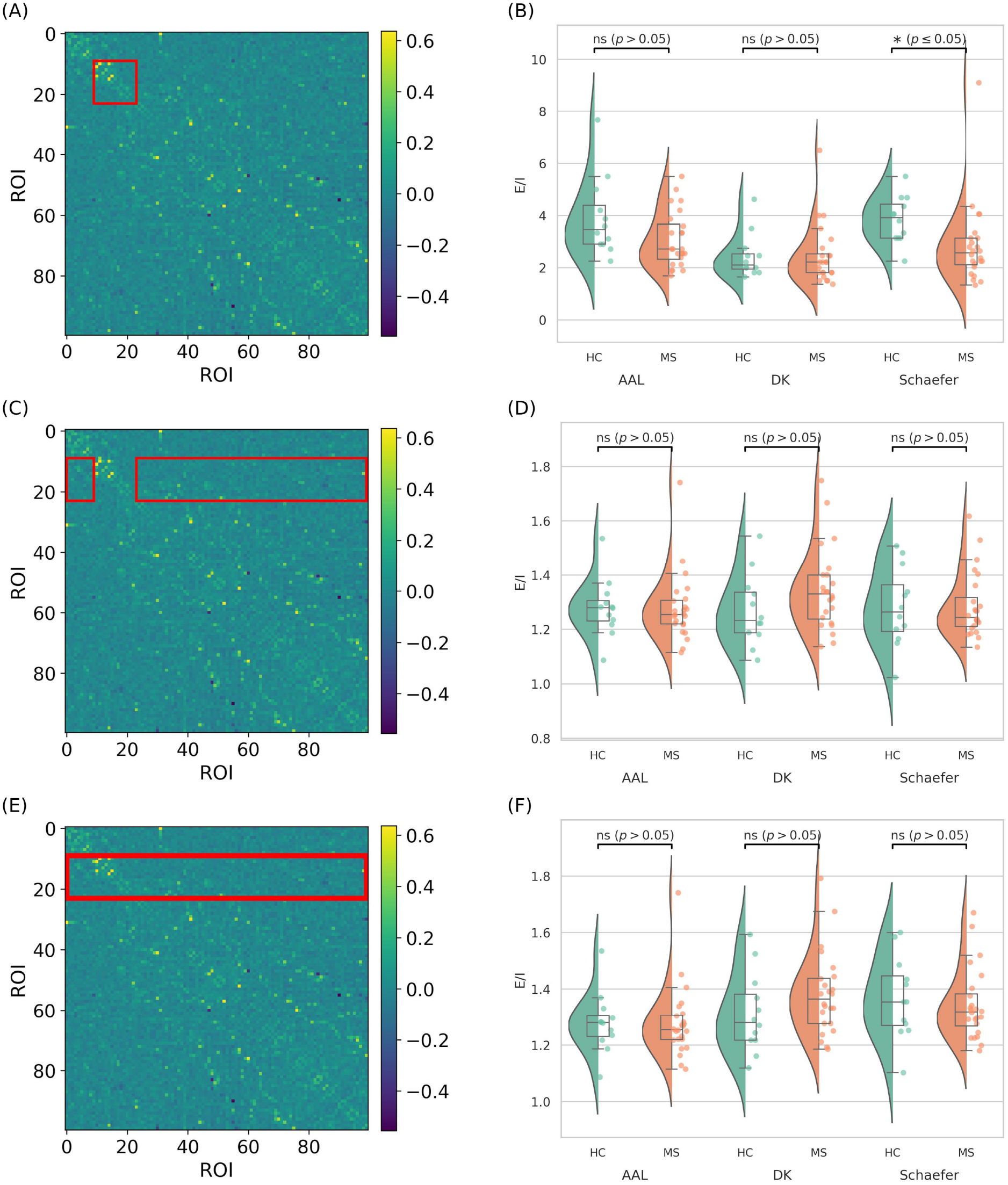
Illustration of the ROIs considered for the E/I calculation for the somatomotor network and statistical distribution of the computed metric between HC and pwMS group. We show the points considered on the rsSC, to calculate E/I in the somatomotor network for points that are (A) intra-network, (C) inter-network, and (E) both inter- and intra-network. On the side, we show the corresponding distribution of EIB values for: (B) ROIs within the somatomotor network, (D) inter-network connections involving the somatomotor network, and (F) both intra- and inter-network ROIs. Each analysis includes three brain atlases: Automated Anatomical Labeling (AAL), Desikan–Killiany (DK), and Schaefer. The Schaefer atlas shows significant differences (*p* ≤ 0.05) after FDR correction for intra-network ROIs. We do not detect significant differences between healthy controls (HC) and people with multiple sclerosis (pwMS) when including inter-network connections.

**Figure 4.**
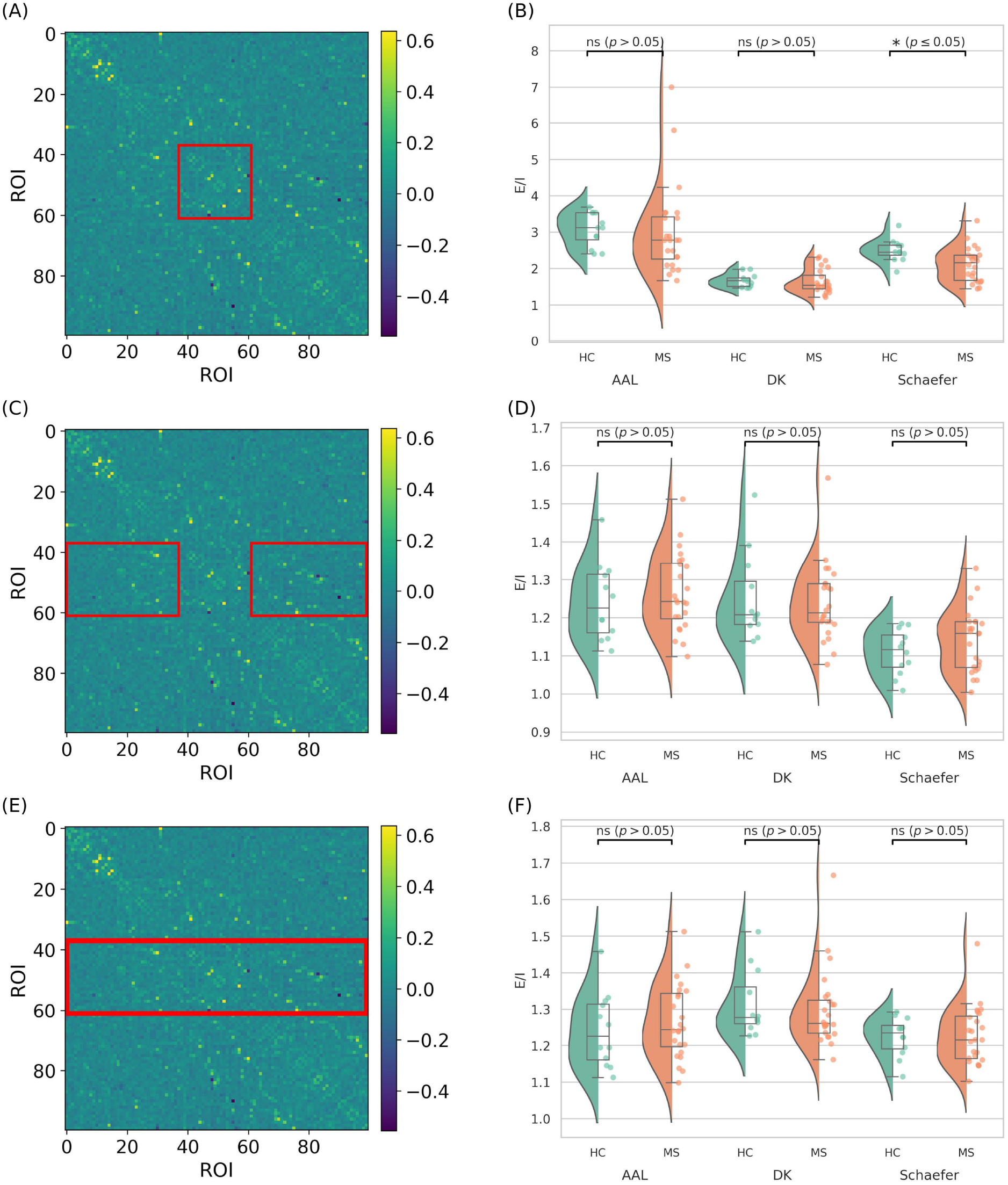
Illustration of the ROIs considered for the E/I calculation for the DMN and statistical distribution of the computed metric between HC and pwMS group. We show the points considered on the rsSC, to calculate E/I in the DMN network for points that are (A) intra-network, (C) inter-network, and (E) both inter- and intra-network. On the side, we show the corresponding distribution of EIB values for: (B) ROIs within the DMN, (D) inter-network connections involving the DMN, and (F) both intra- and inter-network ROIs. Each analysis includes three brain atlases: Automated Anatomical Labeling (AAL), Desikan–Killiany (DK), and Schaefer. The Schaefer atlas shows significant differences (*p* ≤ 0.05) after FDR correction for intra-network ROIs. We do not detect significant differences between healthy controls (HC) and people with multiple sclerosis (pwMS) when including inter-network connections.

We then examined inter-network connections, defined as connections where only one ROI belongs to the network of interest (see **Figures** 3(C) and 4(C) respectively). In this case, the violin plots (see **Figures** 3(D) and 4(D) respectively) showed no significant differences, regardless of the atlas used. Finally, we analyzed the E/I values across all connections that included at least one node within the network **Figures** 3(E) and 4(E). This analysis did not show significant differences between networks or atlases (see **Figures** 3(F) and 4(F) respectively).

### 3.3 Correlation between E/I balance and clinical parameters in MS

We also examined whether intra-network E/I values in the DMN and somatomotor network were associated with cognitive and motor impairment. For the DMN, we assessed correlations with the FSMC–cognitive, COWAT, SDMT, and BVMT, computing Pearson coefficients after removing outliers. For the somatomotor network, we tested correlations with the FSMC–motor and EDSS. As shown in **Figure** 5, we found a significant negative correlation between FSMC–motor scores and somatomotor E/I balance. The correlation between FSMC–cognitive scores and DMN E/I values was also negative and approached significance (*p* = 0.07). No other correlations were significant. Full details are provided in section 4 of the Supplementary Material.

**Figure 5.**
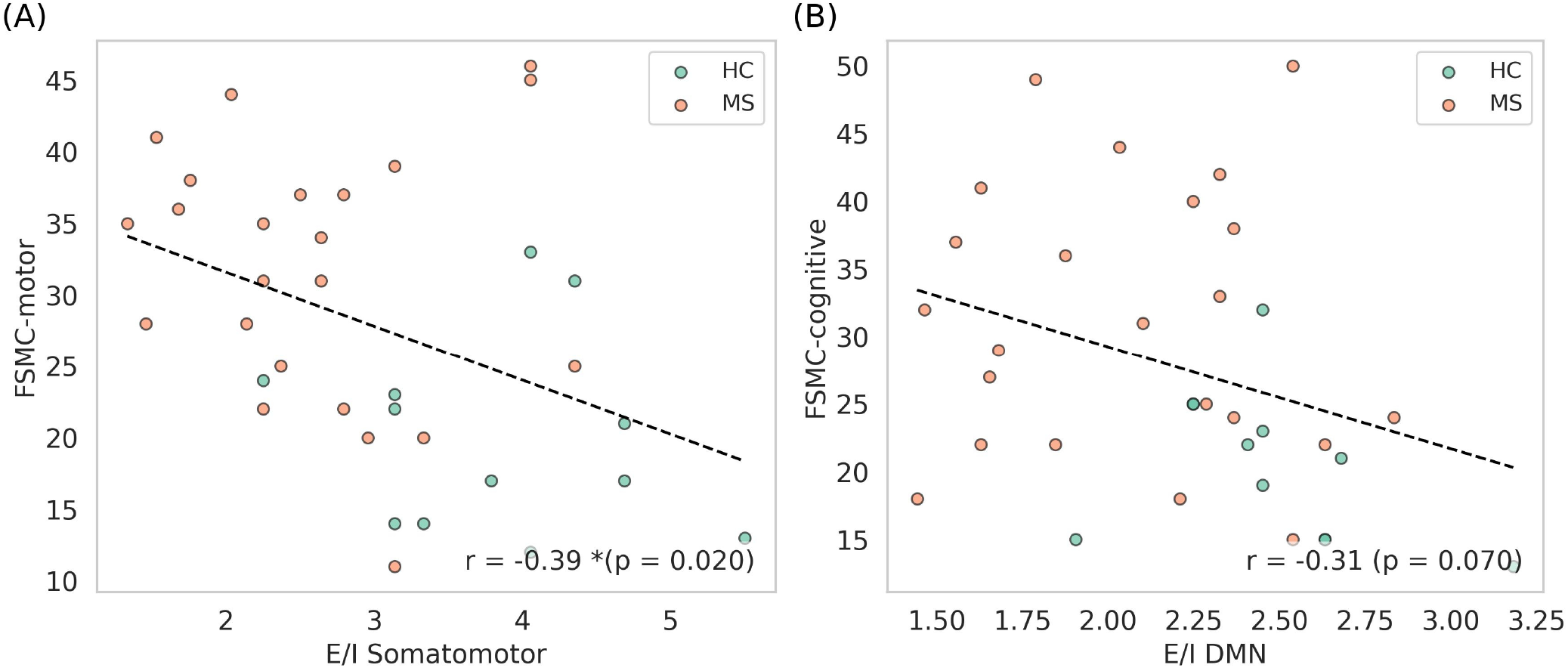
Scatter plots of the motor and cognitive components of the FSMC scores, plotted for each inferred E/I balance. In **(A)**, we show the motor component of the FSMC for each subject, with the E/I balance values from the intra-somatomotor network plotted on the horizontal axis. In **(B)**, a similar plot is shown using the cognitive component of the FSMC score and the E/I balance from the intra-DMN. In this visualization, outliers have been removed. The Pearson correlation coefficient for each plot is shown in the bottom right corner of the respective panel. The motor component of the FSMC shows a significant negative correlation with the E/I balance in the intra-somatomotor network.

## 4 DISCUSSION

In this study, we applied a novel way to assess E/I balance in pwMS. The E/I balance is difficult to assess in vivo, and most studies on the E/I balance in MS rely on electroencephalography or magnetoencephalography. However, most neurofunctional studies in MS rely on fMRI. Here, we apply a recently proposed novel measure of E/I balance based on the so-called resting-state structural connectivity matrix. Similar to Fortel et al. [23], we defined the E/I balance as the ratio of positive (excitatory) to negative (inhibitory) connections in the rsSC matrix and studied it from a global and local perspective. In particular, we examined intra-network, inter-network, and combined connectivity within the somatomotor and DMN networks. Brain regions were defined using the functionally derived Schaefer atlas, which aligns with resting-state networks. For comparison, we also employed the Desikan–Killiany and AAL structural atlases to assess the influence of atlas choice on the results. Our findings highlight network-specific alterations in the E/I ratio in pwMS. These differences are network-specific and only appear within the DMN and somatomotor network. Precisely, alterations in E/I balance were not apparent inter-network or at a global level. In particular, intra-network DMN E/I exhibited a significant group difference (*p* = 0.048, FDR-corrected; medium effect size = 0.36), with the MS group showing lower E/I values than HC. This finding indicates a diminished excitatory drive or enhanced inhibitory tone during resting-state activity in pwMS. Similarly, our analysis reveals significant differences between HCs and pwMS for E/I intra-somatomotor network (*p* = 0.016, FDR corrected; large effect size = 0.54).

Previous studies examining E/I alterations specifically within the DMN in MS generally point to a loss of inhibitory tone. For example, a magnetic resonance spectroscopy study reported reduced GABA concentrations in DMN regions of patients with relapsing–remitting MS [53], although glutamatergic metabolites were not assessed. Additionally, Broeders et al. [54] observed greater functional instability in cognitively impaired MS patients compared with cognitively preserved individuals. They reported frequent reconfiguration of regions of interest into different resting-state networks, with the DMN showing particularly high levels of reconfiguration, an effect the authors interpreted as reflecting increased disinhibition within the DMN and consequently increased E/I balance. In contrast, our findings suggest lower E/I values within the DMN in MS. Importantly, however, the E/I metric used here does not directly capture microscale neuronal or synaptic alterations [25]. Instead, it reflects large-scale connectivity changes that may arise downstream of cellular-level processes such as synaptic loss or shifts in the excitation–inhibition relationship. Thus, our measure may capture a macroscopic consequence of such processes rather than the processes themselves. Furthermore, we detected significant differences only for connections within the DMN, and not for those projecting outside the network. This could indicate that the effects are specific to the DMN, that they arise from interactions between the DMN and certain other networks, or that a larger sample size may be needed to identify effects involving the DMN and other brain regions.

Several studies of the somatomotor network in MS, primarily in animal models, have reported a pro-excitatory shift. For example, Potter et al. [17] found increased excitatory synapse density and loss of inhibitory interneurons in the primary somatomotor cortex, in experimental autoimmune encephalomyelitis. Other studies have also reported similar findings, see e.g., [55, 56, 57]. However, as for the DMN, our results capture a decrease intra-somatomotor network of E-I, describing a macroscopic effect. The employed method did not capture any significant alteration in E/I balance when including all connections involving the somatomotor network.

Furthermore, we observed all significant findings only when we used the Schaefer atlas. This is important because different atlases vary in how well they capture functional networks. We compared the Schaefer atlas with two structural atlases that researchers often use in fMRI studies and found that the functionally defined atlas detected differences more effectively when precise alignment with RSNs was critical. In contrast, anatomically based atlases such as DK and AAL showed lower sensitivity and often failed to detect network-specific functional changes.

Lastly, we examined the linear correlations between intra-network E/I values in the somatomotor network and DMN, computed using the Schaefer atlas, and clinical disability measures (Supplementary Material, **Figures** S7 and S8). The somatomotor intra-network E/I values showed a significant negative correlation with the FSMC motor component, indicating that lower E/I values are associated with greater motor fatigue.

One limitation of this study is the relatively small sample size, which reduces statistical power and limits generalizability. Future studies should include larger cohorts to validate and extend these findings. Additionally, investigating E/I balance during task-based paradigms, would be interesting, especially in relation to the DMN deactivation during working memory task.

## 5 CONCLUSIONS

This study highlights the potential of resting-state structural connectivity to infer E/I (im)balances in pwMS using MRI data alone. A significant reduction in E/I values was observed within the default mode and somatomotor networks of pwMS, suggesting a shift toward increased inhibitory influence within these systems. Furthermore, this reduction was significantly correlated with motor and cognitive fatigue. This pro-inhibitory alteration may reflect underlying neurophysiological adaptations or compensatory mechanisms associated with disease progression at a macroscopic level. The results suggest that functionally defined atlases might better highlight differences in this type of analysis.

## ACKNOWLEDGMENTS

We gratefully acknowledge the fruitful discussions with Liang Zhan. We wish to thank Johan Baijot for the original versions of both the structural and functional pipeline and for the insightful conversations regarding them. The authors also thank all participants in this study for their commitment and participation.

## DATA AVAILABILITY STATEMENT

Due to privacy regulations in accordance with the General Data Protection Regulation (EU) 2016/679, the MRI data from this study cannot be shared publicly. However, researchers interested in accessing the data for collaborative purposes are encouraged to reach out to the senior authors, Prof. Jeroen Van Schependom and Prof. Guy Nagels. The code to compute the hybrid rsSC connectomes and E/I balance can be found at: https://github.com/iforte2/hybrid-connectome

