## Supplementary material for "Altered excitation-inhibition balance in the somatomotor and default mode network in multiple sclerosis"

#### 1 ALGORITHM FOR OBTAINING THE HYBRID RESTING-STATE STRUCTURAL CONNECTIVITY MATRICES

In this section, we describe the algorithm used to generate the hybrid (or signed) resting-state structural connectomes (rsSC), as presented in [1, 2, 3, 4, 5, 6]. This approach integrates structural connectivity with functional time-series data to construct a connectome that characterizes patterns of neural excitation and inhibition. An energy-based description of neural activity is constructed using the statistical-mechanics Ising model, avoiding reliance on standard BOLD correlation measures. In this framework, each pair of brain regions is represented by a coupling parameter (positive or negative) and corresponding spin states, while recorded functional time series capture the evolving brain states. The coupling parameters are estimated through constrained maximum pseudolikelihood, where the constraint acts as a penalty that scales inferred interactions according to structural connectivity. The sign of each estimated interaction can indicate inhibitory or excitatory influences on the underlying structural network.

The pipeline begins by binarizing the preprocessed and z-scored BOLD time series $x_i$ at each ROI $i$ according to $s_{i,t} = \mathrm{sign}(x_{i,t})$. In this way, a brain region $i$ is considered active if $s_i = +1$ and inactive if $s_i = -1$. This step links the functional time series to the spin states of the Ising model. We write the vector of states at time $t$ as $\mathbf{s} = [s_1, s_2, \ldots, s_k]$, where $k$ is the number of ROIs. The time series is first standardized using $z-$score normalization to achieve zero mean and unit variance. Because the interaction term (denoted as $J_{i,j}$ between two regions should correspond to their structural connectivity derived from diffusion MRI tractography, we impose a constraint on the Hamiltonian:

$$H(\mathbf{s}) = \sum_{i<j} J_{i,j} s_i s_j \tag{S1}$$

such that $|J_{i,j}| \propto W_{i,j}$, where $(W_{i,j})$ denotes the structural connectivity (weights) between each pair of ROIs. For resting-state data, external fields are omitted. This formulation ensures that during pseudolikelihood estimation, the inferred interaction matrix J is guided by structural connectivity, assuming that anatomy shapes the spin dynamics. The optimal J is then obtained by maximizing the pseudolikelihood function. Then, the optimal interaction matrix **J** is obtained by maximizing the probability function $Pr(\mathbf{s})$ of observing the (functional) states s at time $t$. The pseudolikelihood function is defined as:

$$\mathcal{L}_{\text{pseudo}}(\mathbf{J}, \beta) = \prod_{t=1}^{t_{\max}} \prod_{i=1}^{k} \Pr\left(s_i(t) \mid \mathbf{J}, \beta, \mathbf{s}_{-i}(t)\right). \tag{S2}$$

Here, $Pr(\mathbf{s})$ is approximated by the product of the conditional probabilities $\tilde{p} = \Pr\left(s_i(t) \mid \mathbf{J}, \beta, \mathbf{s}_{-i}(t)\right)$ of observing the state $s_i(t)$ given all the other states $\mathbf{s}_{-i}(t)$, the interaction matrix **J** and the variable $\beta$ representing the temperature of the system (from the Ising model). We also note that $t_{max}$ depends on the duration of the fMRI series. The log-pseudolikelihood with this penalty term is:

$$\ell(\mathbf{J}, \beta) = \frac{1}{t_{\max}} \ln \mathcal{L}_{\text{pseudo}}(\mathbf{J}, \beta) - \frac{\lambda}{2} \sum_{i<j} \left(J_{i,j} - \mathrm{sgn}(J_{i,j}) W_{i,j}\right)^2.$$

The pseudolikelihood component expands as follows:

$$\frac{1}{t_{\max}} \ln \mathcal{L}_{\text{pseudo}}(\mathbf{J},\beta) = \frac{1}{t_{\max}} \sum_{t=1}^{t_{\max}} \sum_{i=1}^{N} \ln\left(\frac{\exp(\beta \sum_{k=1}^{N} J_{i,k} s_i(t) s_k(t))}{\exp(\beta \sum_{k=1}^{N} J_{i,k} s_k(t)) + \exp(-\beta \sum_{k=1}^{N} J_{i,k} s_k(t))}\right). \tag{S3}$$

This method relies on the Boltzmann distribution under pseudolikelihood assumptions. The numerator captures the system's energy, while the denominator sums over all possible energy states. With si(t) taking on only two binary values, the denominator consists of just two terms—one for each possible sign. The likelihood expression can therefore be simplified by setting: $C_i(t) = \beta \sum_{m=1}^{k} J_{i,m} s_m(t)$, resulting in:

$$\ell(\mathbf{J},\beta) = \frac{1}{t_{\max}} \sum_{t=1}^{t_{\max}} \sum_{i=1}^{N} C_i(t) s_i(t) - \ln(\exp(C_i(t)) + \exp(-C_i(t))) - $$
$$- \frac{\lambda}{2} \sum_{i<j} (J_{i,j} - \text{sgn}(J_{i,j} W_{i,j}))^2. \tag{S4}$$

After defining $C_i(t) = \beta \sum_{m=1}^{k} J_{i,m} s_m(t)$ and we can formulate the probability distribution of the states using the Boltzmann distribution under pseudolikelihood condition. In this way, we can rewrite Eq. (S3) as:

$$\ell(\mathbf{J}, \beta) = \frac{1}{t_{\max}} \sum_{t=1}^{t_{\max}} \sum_{i=1}^{N} \left( C_i(t) s_i(t) - \ln\left(\exp(C_i(t)) + \exp(-C_i(t))\right)\right)$$
$$- \frac{\lambda}{2} \sum_{i<j} \left(J_{i,j} - \text{sgn}(J_{i,j}) W_{i,j}\right)^2. \tag{S5}$$

Finally, to optimize the matrix $\mathbf{J}$, we apply the gradient ascent method:

$$\frac{\partial \ell}{\partial J_{i,j}} = \frac{1}{t_{\max}} \sum_{t=1}^{t_{\max}} \beta \left\{ s_i(t) s_j(t) - s_j(t) \tanh(C_i(t)) \right\} - \lambda \left(J_{i,j} - \text{sgn}(J_{i,j}) W_{i,j}\right) \tag{S6}$$

$$\propto \frac{1}{t_{\max}} \sum_{t=1}^{t_{\max}} \left\{ s_i(t) s_j(t) - s_j(t) \tanh(C_i(t)) \right\} - A \left(J_{i,j} - \text{sgn}(J_{i,j}) W_{i,j}\right). \tag{S7}$$

In this last equation $A = \lambda/\beta$. The update rule for the interaction matrix is:

$$J_{i,j}^{n+1} = J_{i,j}^{n} + \gamma \left.\frac{\partial \ell}{\partial J_{i,j}}\right|_{n}. \tag{S8}$$

Here, $n$ is the iteration number, and $\gamma$ is the learning rate. The estimated matrix $\mathbf{J}$ represents the resting-state structural connectivity (rsSC), or hybrid connectivity. Next, the pipeline optimizes the parameters $A$ and $\beta$. Two metrics guide this process. The first metric, $r_{\text{structural}}$, is the Pearson correlation between the rsSC and the empirical structural connectivity ($\mathbf{W}$). The second metric, $r_{\text{functional}}$, is the correlation between the empirical functional connectivity and the simulated one. The simulated functional connectivity is obtained by using the rsSC as input to Markov Chain Monte Carlo-based Ising model simulations. The values of $r_{\text{structural}}$ and $r_{\text{functional}}$ are computed for different combinations of $A$ and $\beta$ values. A grid search then identifies the optimal parameters giving max $r_{\text{structural}} + r_{\text{functional}}$. This procedure gives equal weight to the reconstruction of structural and functional.

#### 2 EXAMPLES OF FUNCTIONAL, STRUCTURAL AND HYBRID RESTING-STATE STRUCTURAL CONNECTIVITY MATRICES

In this section, we provide examples of the three types of connectivity matrices (functional, structural and hybrid) that are used in this project. In **Figure** S1, we show the three different connectivity matrices for the three different atlases considered (AAL, DK and Schaefer) for a Healthy Control subject.

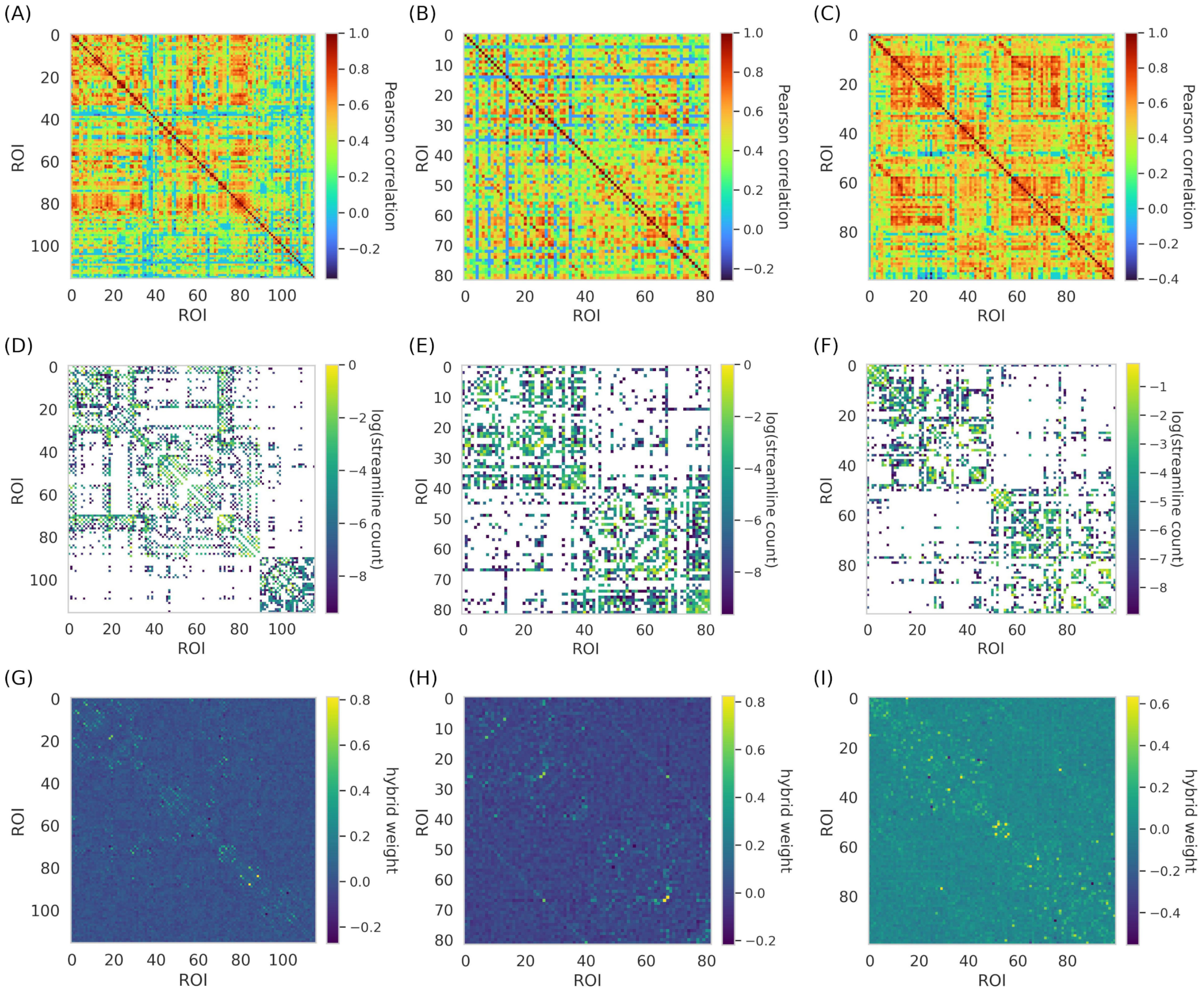


**Figure S1. Functional, structural and rsSC of one HC subject, for every considered atlas.** (A), (B), (C) show the functional connectivity matrices of one healthy control (HC) subject using the AAL, DK, and Schaefer atlases. (D), (E), (F) show the corresponding structural connectivity matrices. We normalize all matrices and plot their logarithmic values for visualization. (G), (H), (I) show the resulting hybrid rsSC matrices for the same atlases.

### 3 RESTING STATE FUNCTIONAL NETWORKS IN THE AAL ATLAS

The structural atlases do not have a direct link to functional networks. Hence, we overlap the AAL with the Yeo7 atlas and we study their overlap [7]. Precisely, we calculate the percentage of voxels in the AAL ROI $i_{\text{AAL}}$ belonging to the Yeo area $i_{\text{Yeo}}$ (**Figure** S2). In **Figure** S4, we study how the Somatomotor networks look on the AAL atlas for the minimum percentage of overlap of 10%, 20%, and 30%. The somatomotor network is composed of a unique 'band' that can be well represented with a percentage of overlap of 30% (**Figure** S4). We note that higher percentages can be investigated as well, taking into account that they would cause the loss of more areas **(Figure** S3. For the DMN, we choose an overlap of 30% as it seems to be well represented **(Figure** S5.

The structural atlases do not directly correspond to functional networks. Therefore, we overlap the AAL atlas with the Yeo7 atlas and analyze their overlap [**?** ]. Specifically, we calculate the percentage of voxels in each AAL ROI $i_{\text{AAL}}$ that belong to each Yeo network $i_{\text{Yeo}}$ (**Figure** S2). In **Figure** S4, we examine how the somatomotor networks are represented on the AAL atlas, using minimum overlap percentages of 10%, 20%, and 30%. The somatomotor network forms a distinct "band" that is most clearly represented with a 30% overlap (**Figure** S4). Higher overlap percentages could also be explored, though they would result in the loss of more areas (**Figure** S3). For the Default Mode Network (DMN), we use a 30% overlap, as it provides a clear representation (**Figure** S5).

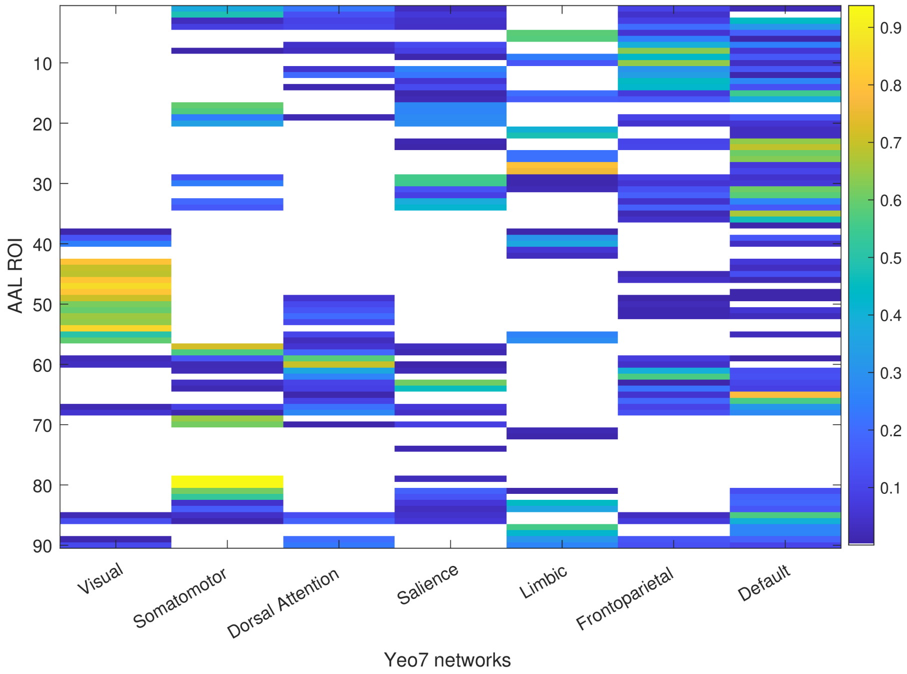


**Figure S2. Overlap between AAL ROIs and Yeo7 Networks.** This analysis reports the percentage of voxels in each AAL ROI that overlap with the seven Yeo functional networks.

Then, we investigate different thresholds of overlap. This means that we study what happens when imposing a minimum percentage of overlap $p_{\text{threshold}}$. In **Figure** S3, we look at how $p_{threshold}$ influences the number of AAL ROIs for the three Yeo networks considered in the previous analysis.

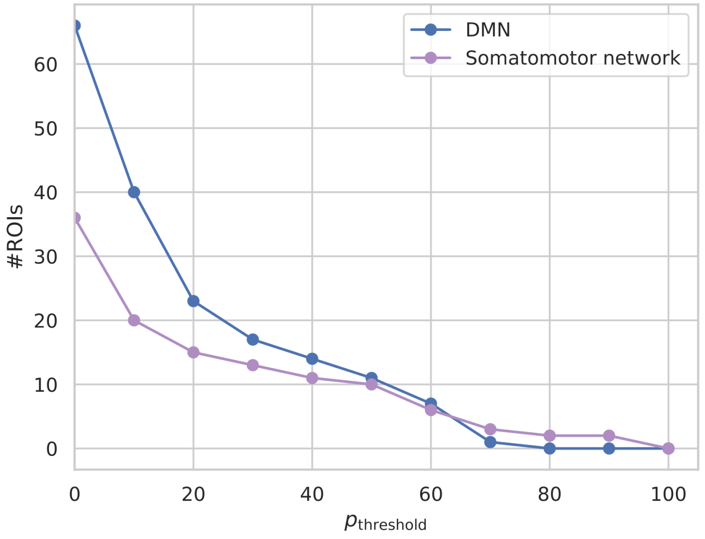


**Figure S3. Analysis of number of AAL ROIs with minimal percentage of overlap with relevant RSNs.** This analysis examines the overlap between AAL ROIs and the DMN and somatomotor network as defined by the Yeo7 atlas. We calculate the percentage of voxels overlapping with the DMN for each AAL ROI. We then count how many ROIs exceed a defined threshold of overlap. This approach helps exclude regions with only marginal involvement in the network.

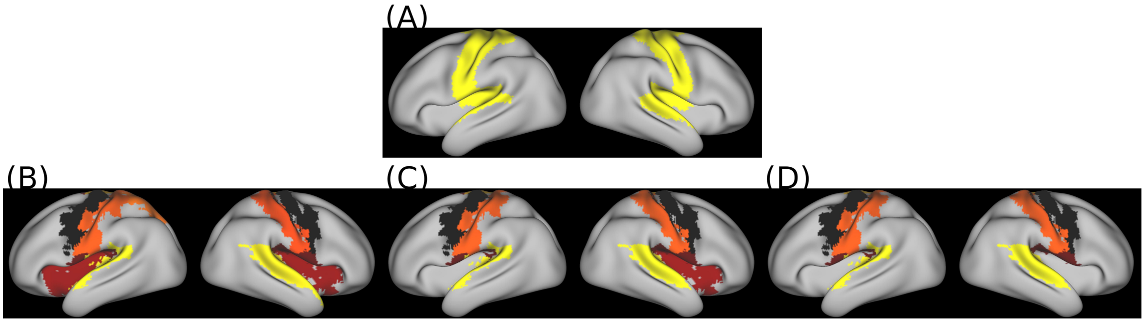


**Figure S4. Somatomotor network mapped onto the AAL atlas, using varying minimal overlap thresholds.** (A) The somatomotor network from the Yeo7 atlas is shown on an inflated brain surface. (B), (C), (D) AAL regions that show at least 10%, 20%, and 30% voxel overlap, respectively, with the somatomotor network from the Yeo7 atlas.

#### 4 LINKING EXCITATION INHIBITION BALANCE TO MULTIPLE SCLEROSIS CLINICAL PARAMETERS

We investigated the association between the inferred intra-network E/I values in the DMN and somatomotor network and clinical measures of cognitive and motor impairment. We quantified these associations using the Pearson correlation coefficient after removing outliers. To remove outliers, we applied a data-driven approach based on the interquartile range. Specifically, we excluded data points whose residuals (to the regression line) fell outside the intervale between $Q1 - 1.5 * IQR$ and $Q3 + 1.5 * IQR$ with $Q1$ and $Q3$ being the first and third quartiles, respectively.

For the DMN, we evaluated correlations with the cognitive subscale of the Fatigue Scale for Motor and Cognitive Functions (FSMC–cognitive), the Controlled Oral Word Association Test (COWAT), the

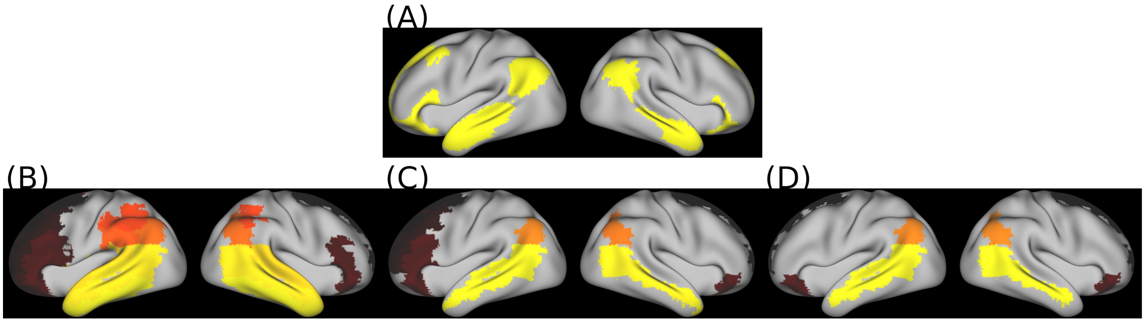


**Figure S5. Default mode network mapped onto the AAL atlas, using varying minimal overlap thresholds.** (A) The DMN from the Yeo7 atlas is shown on an inflated brain surface. (B), (C), (D) AAL regions that show at least 10%, 20%, and 30% voxel overlap, respectively, with the DMN from the Yeo7 atlas.

Symbol Digit Modalities Test (SDMT), and the Brief Visuospatial Memory Test–Revised (BVMT). For visualization, we indicate whether each E/I value belongs to the HC group (green) or the patient group (orange), although the correlation analysis includes all participants. The points indicated in grey (in the correlation with the FsMC-cognitive and BVMT scores) are the outliers identified by our data-driven algorithm. In both cases, the outlier belonged to the MS group. We did not observe any statistically significant correlations.

For the somatomotor network, we examined correlations with the FSMC-motor and the Expanded Disability Status Scale (EDSS). **Figure** S7 presents these results. An outlier belonging to the MS group was also identified for the correlation with the FSMC-motor and EDSS scores. This analysis revealed a statistically significant negative correlation between the inferred E/I values and the FSMC-motor scores.

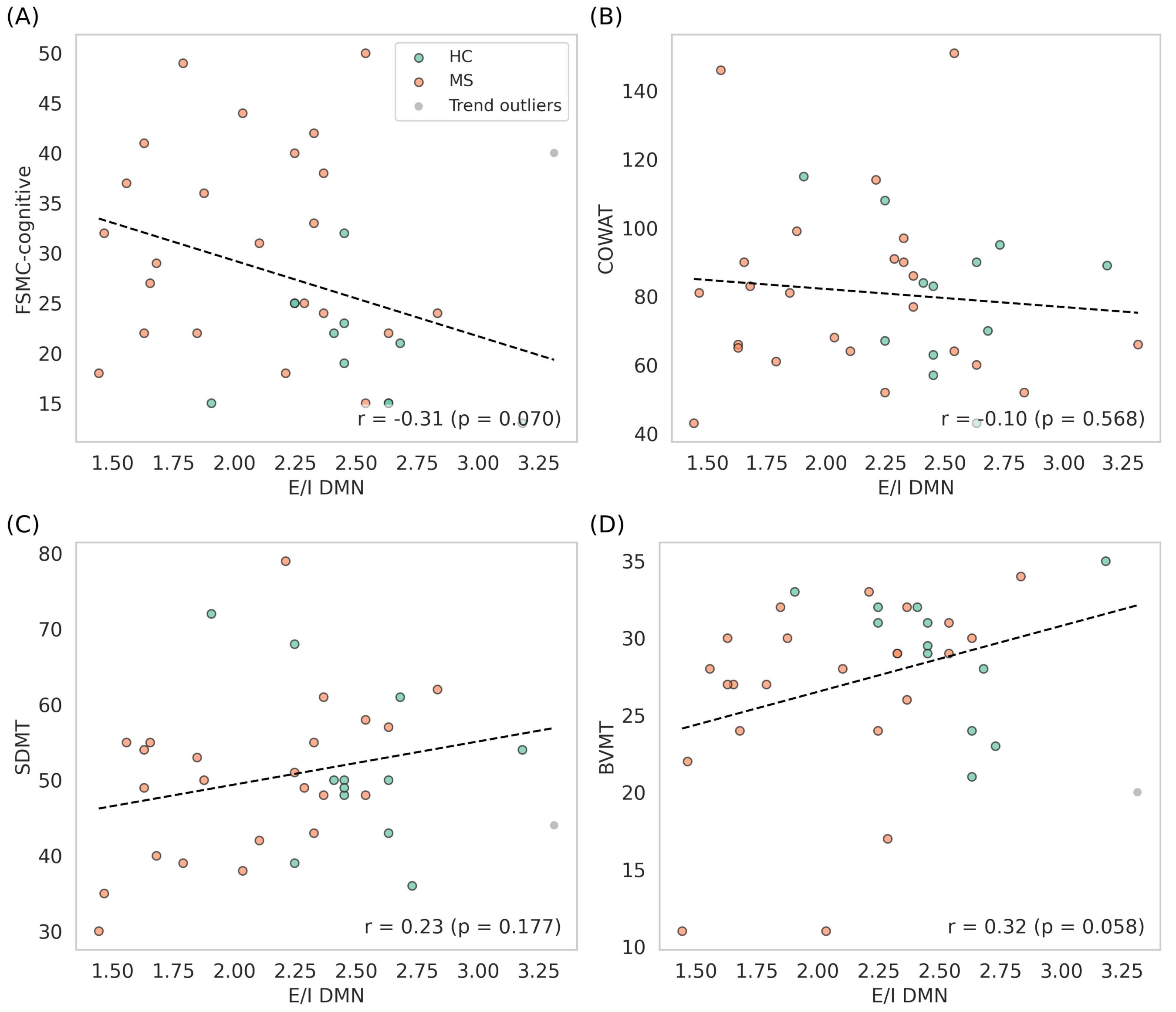


**Figure S6. Correlation analysis between E/I balance in the DMN and cognitive clinical measures.** Pearson correlation coefficient between intra-network E/I values in the DMN and (A) FSMC (cognitive), (B) COWAT, (C) SDMT, and (D) BVMT scores. Grey points indicate outliers removed before the analysis. We found no statistically significant correlations.

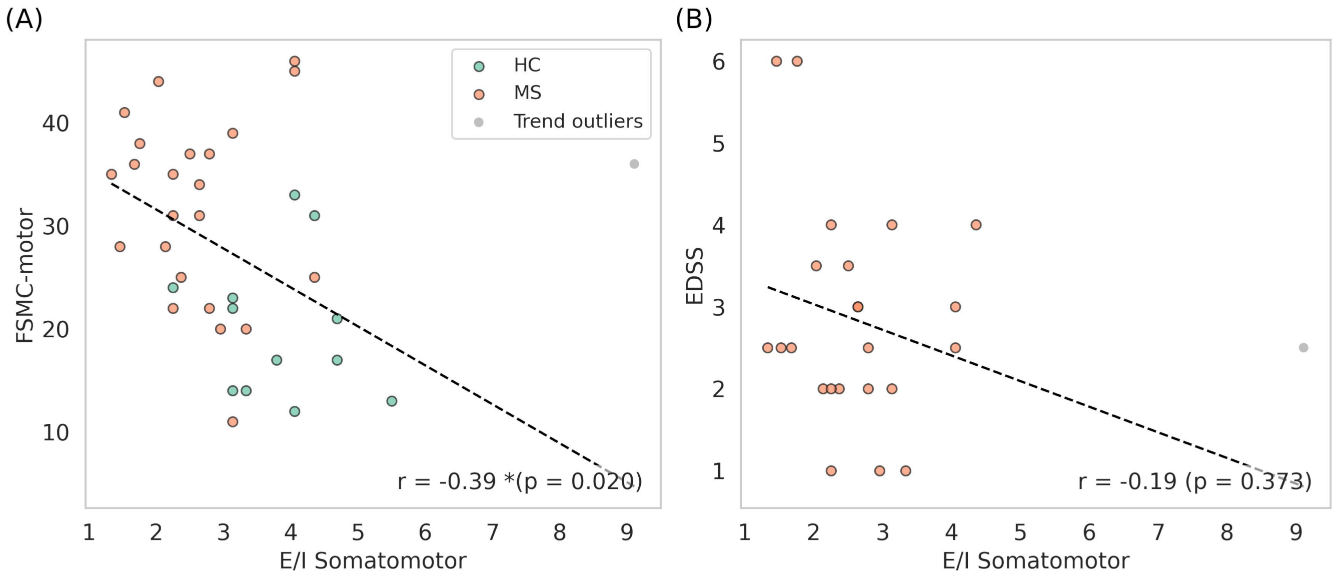


**Figure S7. Correlation analysis between E/I balance in the somatomotor network and motor impairment.** Pearson correlation coefficient between intra-network E/I values in the somatomotor network and (A) FSMC (motor) and (B) EDSS scores. Grey points indicate outliers removed before the analysis. We found a statistically significant negative correlation with the FSMC (motor) score.
